# Considering the methodological limitations and external validity issues of pharmacological drug trials in adult ADHD: An umbrella review (Open Protocol)

**DOI:** 10.1101/2021.12.07.21267067

**Authors:** Kim Boesen, Asger Sand Paludan-Müller, John PA Ioannidis

**Affiliations:** Meta-Research Innovation Center Berlin (METRIC-B), Berlin Institute of Health at Charité, Berlin, Germany; Centre for Evidence-Based Medicine Odense (CEBMO) and Cochrane Denmark, Department of Clinical Research, University of Southern Denmark, JB Winsløwsvej 9b, 3rd Floor, 5000 Odense, Denmark; Open Patient data Exploratory Network (OPEN), Odense University Hospital, Odense, Denmark; Meta-Research Innovation Center at Stanford (METRICS), Stanford University, Stanford, CA, USA; Departments of Medicine, of Epidemiology and Population Health, of Biomedical Data Science, and of Statistics, Stanford University, Stanford, CA, USA

## Abstract

This is a protocol to an umbrella review entitled, ‘Considering the methodological limitations and external validity issues of pharmacological drug trials in adult ADHD: An umbrella review (Open Protocol)’.

## Introduction

Systematic reviews of clinical trials assessing the benefits and harms of the central nervous system (CNS) stimulants methylphenidate^1-3^ and amphetamines^4^ for adults with attention deficit hyperactivity disorder (ADHD) have highlighted various methodological limitations of the evidence base. These limitations include short follow-up, high risk of bias, a lack of patient reported outcomes, and limitations to the trials’ generalisability or ‘external validity’. Several design issues may affect the external validity of psychiatric drug trials, as already highlighted for trials of antidepressants^5^ and antipsychotics.^6^ We would like to showcase three specific designs issue that remain to be characterised for a large body of ADHD drug trials.

### Issue 1: strict inclusion criteria

The first design issue pertains to strict inclusion criteria related to psychiatric comorbidity. Adults diagnosed with ADHD have high rates of psychiatric comorbidity.^7-9^ If randomised trials impose strict exclusion criteria, the tested population may not reflect those treated in a clinical setting.^10^ This is well described in other psychiatric fields like depression trials.^11-13^

### Issue 2: responder selection (‘enriched design’)

The second design issue relates to previous exposure to the same, or a similar, drug. If a clinical trial stipulates to allow previous exposure to the tested drug, or from the same drug class, and selects participants based on the previous treatment response, this is called an ‘enriched design’.^14^ This means that the population is ‘enriched’ in order to amplify potential beneficial signals of effect. Such trial design may be justified in specialties, e.g. oncology, where participant selections may be based on clinical markers, both in the trial and in a clinical setting. In specialties without the possibility of such diagnostics, as psychiatry, any treatment response selection will lead to an overestimation of the beneficial effects and an underestimation of the harms compared to a treatment naïve population.

### Issue 3: withdrawal effects

The third design issue relates to potential confounding by withdrawal effects. Withdrawal of psychotropic drugs, like CNS stimulants such as cocaine, amphetamine, and methamphetamine may lead to acute and protracted withdrawal effects lasting weeks to months.^15-19^ We also consider it plausible that other drug types used for ADHD, e.g. atomoxetine and bupropion, carry a risk of withdrawal effects (see appendix for a detailed argument).

If (a proportion of the) trial participants are already taking ADHD medication, like methylphenidate or atomoxetine, upon enrolment and taper their medication before randomisation, this may introduce withdrawal effects. Participants may experience withdrawal effects during the trial if the taper of concurrent medication is not of sufficient duration. Those randomised to placebo may worsen because of withdrawal effects, whereas those who are randomised to the active drug may have these effects alleviated. This difference between the groups may mistakenly be interpreted as symptom improvement caused by the drug, whereas - in fact - it may have been an iatrogenic artefact.

It will be useful to generate an overarching view on these pertinent limitations based on published systematic reviews. See Table 1 for a proposal to operationalise the categorisation of these trial characteristics.

**Table 1.** Trial design characteristics

|  | **Restricted populations** | **Enriched design** | **Withdrawal effects** |
| --- | --- | --- | --- |
| **Low risk of affecting the external validity** | Participants with relevant psychiatric comorbidity were allowed to participate.<br>Reasonable exclusion criteria would include schizophrenia, mania, and suicidal behavior. | Participants were CNS stimulant treatment naïve. | Tapering of existing CNS stimulant treatment were of sufficient duration to avoid withdrawal effects.<br>We define this as a **minimum of one year** or more without intake of CNS stimulants. |
| **Unclear risk of affecting the external validity** | Insufficient description of in and exclusion criteria. | Insufficient description of in and exclusion criteria. | The tapering period may not be of sufficient duration.<br>We define this period as **between 6 to 12 months**. |
| **High risk of affecting the external validity** | Participants with relevant psychiatric comorbidity were excluded from the trials.<br>These comorbidities include depression, anxiety, and personality disorders. | Participants with previous exposure to the tested drug were allowed to participate;<br>and/or the participants were selected based on their treatment response. | The tapering period was insufficient to avoid withdrawal effects.<br>We define this period as **six months or less**. |

## Methods

This is a protocol to an umbrella review aiming to synthesise the evidence gathered in different systematic reviews. The results should be reported according to the PRISMA guideline.^20^

### Research objective

To assess pharmacological drug trials in adults with ADHD with a focus on trial design characteristics that impact the external validity.

### Project type

This is a review of systematic reviews, also called an umbrella review.

### Eligibility criteria for systematic reviews

1. Study type: Systematic reviews
  - *Published in the Cochrane Database of Systematic Reviews*,^21^ *with a pre-registered protocol specifying outcomes and methodology*.
  - *We will not search for, or include, reviews published outside the Cochrane Library*.
2. Population: Adults diagnosed with ADHD.
3. Intervention: Any pharmacological drug used for ADHD treatment
  - *E.g. methylphenidate, amphetamines, atomoxetine, and bupropion, in any dose and in any formulation, e.g. immediate and extended-release formulations*.
4. Comparison: Placebo.
5. Outcomes: Benefits and harms, no restrictions.

### Searching for systematic reviews

We will search the Cochrane Database of Systematic Reviews^21^ for published systematic reviews on pharmacological interventions for adult ADHD. See preliminary search results in Table 2.

**Table 2.** Eligible reviews

| Review | Status | Drug | External validity issues assessed | Trials (n) | Population (n) |
| --- | --- | --- | --- | --- | --- |
| Boesen et al. (2017)<sup>3, 25</sup> | Protocol<sup>a</sup> | Extended-release methylphenidate | Yes | 25 | 5066 |
| Venables et al. (2017)<sup>39</sup> | Withdrawn (only protocol was published)<sup>b</sup> | Alpha<sub>2</sub> adrenergic agonists | No | N/A | N/A |
| De Crescenzo (2018)<sup>26</sup> | Protocol | Noradrenergic reuptake inhibitors (mainly atomoxetine) | Yes | N/A | N/A |
| Verbeeck et al. (2017)<sup>24</sup> | Final review | Bupropion | No | 6 | 438 |
| Castells et al. (2018)<sup>4</sup> | Final review | Amphetamines | No | 19 | 2521 |
| Candido et al. (2021)<sup>2</sup> | Final review | Immediate-release methylphenidate | No | 10 | 497 |
| **Total** |  |  |  | 60 | 8522 |

<sup>a)</sup> The review is not yet published. We are authoring it and have the final results.
<sup>b)</sup> The protocol to this review has been withdrawn and will not be included in our analysis.

### Outcomes

We will extract the following information from the clinical trials included in the reviews:

1. ***Basic trial information***
  a. *Trial IDs, period of conduct, trial registry number, and funding (industry/public/both)*.
2. ***Sample size***
3. ***Trial duration***
4. ***Risk of bias***
  a. As judged by the authors in the systematic reviews.
5. ***Patient reported outcomes***
  a. Quality of life (self rated).
  b. Functional outcomes, i.e. any outcome measuring functional capacity. Rating scales, like Sheenan Disability Scale,^22^ will not be included.
6. ***Trial design 1: Restricted trial population concerning psychiatric comorbidity***
  a. We will assess if the trials imposed strict exclusion criteria related to psychiatric morbidity other than ADHD.^23^
  b. See criteria in Table 1.
7. ***Trial design 2: ‘Enriched design’***
  a. We will assess if the individual trials employed an ‘enriched design’.^23^
  b. See criteria in Table 1.
8. ***Trial design 3: Withdrawal effects***
  a. We will assess whether the individual trials were at risk of introducing ‘withdrawal effects’ to those randomised to placebo.^23^
  b. See criteria in Table 1.

### Analyses

1. ***Sample size versus trial duration*** We will depict the accumulated sample size over trial duration. As an example, see reference 5 (supplement 2, figure 1).^5^
2. ***Summary of risk of bias assessment*** We will summarise domains rated as ‘unclear’ and ‘high risk’. We assume all reviews have used the original Cochrane Risk of Bias tool.
3. ***Effect sizes of patient reported outcomes*** Reported as mean differences or standardised mean differences. We will prefer to report mean differences. We will summarise the results, if feasible, in random effects meta-analyses using inverse variance weighting.
4. ***Prevalence of trial design issues*** We will report how many trials employ ‘enriched design’; have strict exclusion criteria; and were at high risk of ‘withdrawal effects’.
5. ***Impact of trial design characteristics on patient reported outcomes*** We will conduct subgroup analyses to assess differences in reported effect sizes depending on the prevalence of the three design issues, i.e. trials with ‘high risk’ of restricted populations compared to trials with a ‘low risk’; trials with a ‘high risk’ of ‘enriched design’ compared to trials with ‘low risk’; and trials with a ‘high risk’ of withdrawal effects compared to trials with ‘low risk’.
6. ***Prevalence of conflicted trials*** We will report the proportion of industry-sponsored trials and publicly funded trials with industry-involvement (e.g. as declared on trial registries or as acknowledgements in published papers).

### Data extraction

According to our preliminary search (Table 2), we identified three^2, 4, 24^ published reviews and protocols for two^25, 26^ reviews meeting our eligibility criteria. We are authoring one of reviews that have not been published,^3, 25^ and we will correspond with the author group of the other protocol^26^ regarding access to the review data.

Two authors should independently extract outcome data from the included reviews and arbiter with a third author, if necessary. Information related to the three trial design issues are reported systematically only in the systematic reviews of extended-release methylphenidate^25^ and atomoxetine,^26^ therefore this information should be extracted from the other reviews’ included trials manually.

## Discussion

To our knowledge, this will be the first umbrella review on the methodological limitations of drug trials in adult ADHD. We believe this umbrella review will add important insight to frequent methodological limitations and showcase which domains that ought to be improved in future trials of ADHD medications. It may likely guide funders and drug regulatory agencies on how new drug trials should be designed.

### Limitations

There are several limitations to this project. First, we propose to not search for reviews published outside the Cochrane Library. We do this to reduce the anticipated workload and to mitigate heterogeneity between the included systematic reviews. The two most recently published non-Cochrane systematic reviews on ADHD medications,^27, 28^ did not assess characteristics related to the external validity, and one of them^27^ did not assess functional outcomes. We judge that the added benefit of including such reviews is limited.

Secondly, we plan to extract data from the individual trials on design characteristics (since we expect this information to be adequately reported in published reports), whereas we do not plan to extract outcome data on patient reported outcomes. To thoroughly do this, it would require searching for unpublished data, regulatory databases, and clinical trial registries. Such efforts may be worthwhile to pursue in separate projects.

Thirdly, our categorisations of the trial design characteristics – especially our arbitrary thresholds for withdrawal duration - are definitely open to discussion. The evidence on CNS stimulant withdrawal effects^15-19^ and the duration of withdrawal symptoms is weak, especially for methylphenidate, and even weaker for drugs like atomoxetine. It seems to be an understudied field of research, which is one of the main reasons we would like to describe their potential occurrence in these trials.

Finally, we risk committing an ecological fallacy by conducting subgroup analyses to test the impact of trial design issues on patient reported outcomes using aggregate group level data rather than using individual patient-level data. However, since this is the first review to assess the frequency of these design issues, we feel obligated to assess their impact, bearing in mind that the analyses are vulnerable to ecological artefacts.

### The ‘Open Protocol’ Framework

This is an ‘Open Protocol’ meaning that it has not been assigned a first author to lead the project. We encourage anyone to contact us if they are interested in working on this project. The protocol will remain open to changes and adjustments until data extraction begins. We will update the ‘Version history’ once the protocol gets assigned a lead author or upon changes to the methodology.

## Data Availability

The final project should be made available as a pre-print and its full dataset made available as a collated datafile assigned with a separate DOI on a suitable repository, like the Open Science Framework or Zenodo.

## Acknowledgements

None

## Version history

**Version 1 (Nov 2021)**

- ***Date*** Submitted to MedRxiv on 30 Nov 2021.
- ***Changes compared to previous version*** None. First version

## Appendix Risk of withdrawal effects

### Atomoxetine

Industry sponsored research^29, 30^ have reported that the norepinephrine reuptake inhibitor atomoxetine carries no risk of withdrawal effects. However, these studies were designed to select participants who tolerated well the drug over an extensive time, e.g. by using the randomised withdrawal design, which may likely have reduced the risk of adverse effects.^31^ To more reliably assess atomoxetine’s withdrawal profile one would need differently designed trials - conducted by others than the marketing holder. We consider it plausible that abrupt stop of atomoxetine, as for other psychotropic drugs,^32, 33^ may cause withdrawal effects.

### Bupropion

Some case reports^34-36^ have described withdrawal effects following use of the norepinephrine dopamine reuptake inhibitor bupropion. As for other antidepressants^32, 33, 37, 38^ it seems plausible that withdrawal effects may occur upon bupropion discontinuation.

### Preliminary search of eligible systematic reviews

Date: 27 Sep 2021.

Search strategy: “Attention deficit hyperactivity disorder”.

Hits: ‘Reviews’ (23), ‘Protocols’ (8). We did not assess ‘Trials’ (4028), ‘Editorials’ (0), ‘Special Collections’ (0), or ‘Clinical Answers’ (5).

Eligible hits: 6.

